# Nurses in Advanced Roles as a strategy for equitable access to health care in the WHO Western Pacific Region: A mixed methods study

**DOI:** 10.1101/2020.05.18.20105056

**Authors:** Sue Kim, Tae Wha Lee, Gwang Suk Kim, Eunhee Cho, Yeonsoo Jang, Seoyoung Baek, David Lindsay, Sally Wai-Chi Chan, Regina Lai Tong Lee, Aimin Guo, Frances Kam Yuet Wong, Doris Yu, Sek Ying Chair, Yoko Shimpuku, Sonoe Mashino, Anecita Gigi Lim, Sheila Bonito, Michele Rumsey, Amanda Neill, Indrajit Hazarika, Mona Choi

## Abstract

**Purpose:** This study investigated the current responsibilities of nurses in advanced roles (NARs), future healthcare needs, and the implications of these components for professional development of nurses.

**Design:** This study employed a descriptive survey on the current status of NARs in the Western Pacific region (WPR), followed by a Delphi survey and exploratory interviews. Experts from WPR countries who were individuals with recognized national expertise on NARs from clinical, academic, and/or government-related backgrounds were invited to participate in this study from December 2017 to December 2018.

**Methods:** Fifteen experts from ten countries provided descriptive data on the current status of NARs in the WPR via email. The data were used to grasp the spectrum of NAR and construct a working definition of NARs. This formed the basis for the Delphi survey, in which 27 experts from 14 countries completed the following three sections via email: Areas in need of NARs; Strengths, Weakness, Opportunities, and Threats analysis; and the role of NARs in promoting universal health coverage. Descriptive statistics were used for the survey and Delphi survey. For the exploratory interviews, semi-structured individual interviews were conducted with 19 key informants from 12 countries. Content analysis was performed for interview data. Delphi and interview findings were integrated in the final stage.

**Findings:** Thirty-seven roles were identified and categorized according to the regulations for the specific roles. Emergency care, critical care, elderly health, child health, and rural/remote communities were identified as fields with particular need for NARs. Providing effective services, influencing government leadership, and advocating for health system sustainability were deemed necessary for NAR to improve equitable healthcare access.

**Conclusions:** Demand for NARs is high in the WPR and we presented 15 items across five core strategic areas within the nursing community to enhance NAR development. Governmental-level recommendations include establishing legislative protection, remuneration, supportive channels, and conducting national needs assessments.

**Clinical Relevance:** NARs are not limited to clinical tasks within the hospital but are poised for active participation in primary healthcare, education/teaching, professional leadership, quality management, and research.

## Introduction

The changing demographic and epidemiological trends in countries of the World Health Organisation (WHO) Western Pacific region (WPR) have resulted in a shift in population healthcare needs, creating an increased demand for health services. Regardless of socio-economic development levels, there is a growing global recognition of the importance of the health worker’s role in promoting equitable access to quality health services. In most countries, nurses represent by far the largest part of the healthcare workforce and are critical to both the universal health coverage (UHC) (WHO, 2016) and Sustainable Development Goal agendas (United Nations, 2015).

Within the nursing workforce, nurses in advanced roles (NARs) have evolved to provide high quality care, especially at the primary care level. NARs are often regarded as similar to nurse practitioners (NP), advanced practice nurses (APN), or midwives. The International Council of Nurses (ICN) defines a NP/APN as “a registered nurse who has acquired the expert knowledge base, complex decision-making skills, and clinical competencies for expanded practice, the characteristics of which are shaped by the context and/or country in which s/he is credentialed to practice,” and specifies that a master’s degree is recommended for entry level (ICN, 2008). These titles and subsequent roles have been embraced and developed in Northern America and other high-income countries such as the UK, the Netherlands, Finland, etc., with legislative protection and graduate-level education taking root (Keeling, 2015; Maier, Batenburg, Birch, Zander, Elliott, & Busse, 2018). In the US, the NP/APN model of clinical excellence recently has been extended to advanced degrees such as the Doctor of Nursing Practice (American Association of Colleges of Nursing, 2015). The International Confederation of Midwives (ICM) defines a midwife as a person who has successfully completed a midwifery education program that is based on the ICM Essential Competencies for Basic Midwifery Practice and the framework of the ICM Global Standards for Midwifery Education (ICM, 2017).

There is a sizeable body of empirical literature (Cashin et al, 2015; Hanson & Hamric 2003; Nardi & Diallo, 2014; Wysocki, Underwood, & Kelly-Weeder, 2015) supporting the unique contributions of NP/APN and midwives. While these titles are widely known and specific to country context, they have not been sufficient nor perhaps relevant to countries with different health systems, resources, and historical contexts. Despite the fact that there are numerous nurses who take on a variety of roles and responsibilities extending beyond their basic training and contributing considerably to healthcare throughout the world, NARs have lacked consistency in nomenclature, scope of practice, role-related regulation, qualification, and educational systems. There is limited empirical literature aside from the aforementioned countries where NP/APN are well established. Furthermore, there has been a lack of concerted effort to examine the status and spectrum of NAR and identify potential contributions, especially in relation to the five essential health system attributes required to achieve UHC —quality, efficiency, equity, accountability, and sustainability and resilience (WHO, 2016).

Thus, the purpose of this study was to examine the current roles of NARs and their contributions to health services in WPR countries, as well as implications for the professional development of NAR to meet future healthcare needs at the national and regional levels. The specific objectives were to 1) identify the current status of NARs in the WPR (e.g. functions, scope, competencies, educational standards, credentialing, and regulation); 2) assess how NARs might be able to improve equitable access to quality healthcare, including the identification of key barriers and challenges; and 3) identify the role of NARs in addressing future healthcare needs, including recommendations on their contributions and roles in the healthcare system.

The WPR includes almost 1.9 billion people in 37 countries and areas (WHO WPR Office, 2020). This multi-country study was led by the WHO Collaborating Centre (WHO CC) of the College of Nursing at Yonsei University, in collaboration with other WHO CCs and key stakeholders from participating countries.

## Methods

The working group of the WHO CC of the College of Nursing, Yonsei University, Korea, held several meetings to plan and form the NAR Study Group to conduct this multi-country study from April to June, 2017. The NAR Study Group was formed using a previously existing network of nursing and midwifery related WHO CCs and finally consisted of representatives from 13 institutions in 8 countries.

This study was conducted in three phases: a descriptive survey, a Delphi survey, and exploratory interviews phases, followed by analysis and integration of data. The study period was December 2017 – December 2018. The approval from the Institutional Review Board was obtained from Yonsei University (Y-2017-0076) and written consent was obtained for all participating experts.

Participants for each phase were recruited using a purposive sampling approach to invite experts who were identified by the NAR Study Group, from WHO CCs as well as from six additional countries where nursing-related CCs did not exist but had NARs placed in nursing profession (Table 1).

### Phase 1: Descriptive Survey

The researchers of the NAR Study Group identified one to three experts from the region, i.e., individuals with recognized national expertise on NARs from clinical, academic, and/or government-related backgrounds. Once they were identified, the Yonsei working group invited the experts to provide descriptive data on the status of NARs within their own country.

After obtaining informed consents, 15 experts from ten countries participated in an email survey consisting of two parts: First, the status of NARs in their countries, e.g., role-related regulations, credentialing, and education preparation. These items were open-ended questions so that respondents could answer according to NAR categories that exist in each country. The second part focused on NARs’ specific roles and responsibilities in 6 areas with 29 items derived from the literature; clinical/technical tasks (11 items), primary care (8 items), education and teaching (4 items), professional leadership (2 items), quality management (2 items), and research (2 items). The email survey was conducted from December 2017 – March 2018. Reminders were sent up to three times via email to ensure responses. Based on this expert input, a working definition of NARs was developed to guide the subsequent phases of the study.

### Phase 2: Delphi method

With a similar sampling approach, the NAR Study Group identified 27 experts from 14 countries for the Delphi phase, which was implemented in three rounds. Experts who met the same selection criteria for Phase 1, preferably experts who had not participated in the previous phase were invited. Those who consented completed the Delphi survey via email from April – July 2018. The Delphi method (Hsu & Sandford, 2007) was employed to identify consensus among experts on how NAR roles might be able to improve equitable access to quality healthcare, including identification of key barriers/challenges and need for the NAR in achieving UHC. The first Delphi survey round asked respondents to rate the status of NARs within their country. Rounds 2 and 3 focused on rating how NAR could contribute to improving equitable access to quality healthcare within the WPR.

The questionnaire consisted of three parts: 1) Areas in need of NARs, which was divided into four sub-domains (field-specific domain, client group-specific domain, area-specific domain, and professional role-specific domain). Twenty-six items were rated on a five-point Likert scale (1 = *not required*, to 5 = *essential function for NARs)*. 2) In part 2, positive and negative influences in developing and establishing advanced nursing roles were assessed using the SWOT (Strengths, Weakness, Opportunities, and Threats) format. Thirty-eight items were rated on a five-point Likert scale (1 = *strongly disagree*, to 5 = *strongly agree)* as well as “not applicable.” 3) In part 3, the role of NARs in the development of UHC was assessed according to the WHO domains (WHO, 2016). Respondents rated the priority for the five following subcategories: quality, efficiency, equity, accountability, and sustainability/resilience. These 15 items were also rated on a five-point Likert scale (1 = *low priority*, to 5 = *high priority)*.

Descriptive statistics were used to describe the Delphi data in each round. After analysing the data collected in round 1, the questionnaire was slightly revised for use in round 2, by deleting four items with low scores. While the same questionnaire was used for rounds 2 and 3, in the third round, the mean and range of round 2 responses were presented to the experts as reference data, and they were asked to answer considering this information or write down the reasons if their chosen response would be out of the range.

### Phase 3: Exploratory interviews

Semi-structured individual interviews with 19 key informants from 12 countries, were conducted to explore and understand how NARs improve access to health services. Furthermore, these interviews served to solicit recommendations for NARs across various health systems to improve equitable access to healthcare in the WPR. This phase was conducted from July – September 2018.

Key informants for the interviews included healthcare professionals (e.g. nurses, physicians), leaders (e.g. nursing leaders, policy makers), regional experts, and members of related networks, such as the South Pacific Chief Nursing and Midwifery Officers Alliance. The NAR Study Group members conducted face-to-face or telephone interviews in the language of the informant to obtain rich and accurate data, and an English summary for each interview was shared among the Group to facilitate discussion and content analysis.

### Phase 4: Analyses and Integration of Phases 2 and 3

Descriptive statistics were calculated for the data from the descriptive survey (phase 1) and Delphi (phase 2). Content analysis was done for the qualitative data from phase 3. All data were analysed by the Yonsei working group over 14 research meetings and reviewed by and revised with input from the larger Study Group. For consistent communication, process monitoring, discussion, and verification of the analysis process, teleconferences with the Study Group were conducted multiple times and supplemented with email discussion. We discussed the specific methods of the study, and monitored the process of written consent from the research partners at every phase. After setting the direction of analysis, the working group analysed the data and wrote up the results for each phase. Further verification of the data and findings were also shared in face-to-face meetings with some research partners during international conferences and visits. Finally, the results of the Delphi and exploratory interview phases were integrated to derive strategies and recommendations for developing and promoting the role of NARs in the WPR.

## Findings

### Phase 1: Status of health systems and NARs in the WPR

In the current study, the term “NAR” was intended to be used very broadly to describe nurses undertaking roles that are beyond the basic level of nursing. We identified 37 scopes of the NARs role from ten countries and categorized the findings according to whether role-related regulations existed. In cases of regulation, roles were classified as to whether they were consistent with the definitions put forth by the ICN NP/APN network (ICN, 2008).

We found that the functions of NARs were not limited to clinical tasks within the hospital but also included primary healthcare, education/teaching, professional leadership, quality management, and research. Even though the roles of NARs did not have regulatory standards in some countries, they were described in standards by nursing and professional associations, job descriptions, and/or training standards by educational institutions. The NARs who satisfied regulatory requirements laid out by the ICN were classified as NPs in Australia, New Zealand, and Fiji, whereas they were classified as specialist nurses or public health officers in the Solomon Islands. There were also legally stipulated roles although these did not necessarily meet the definition put forth by the ICN. These were APN (midwife specialist) role or midwife consultants in Hong Kong; public health nurses or midwives in Japan; specialized nurses (home healthcare) or public officials exclusively responsible for public healthcare services in South Korea; clinical nurse specialists with prescribing rights in New Zealand; and in the Philippines, clinical nurse specialists based in hospitals (e.g. enterostomy nurses, geriatric nurses) or potential NPs in communities (e.g., nurse-midwives, nurse diabetes educators). The minimum level of education required differed depending on the role and country. The education level ranged from holding an advanced diploma to a master’s degree.

Based on the data, the following working definition of NARs was formulated for use in the current study:

> *“A registered nurse who practices beyond basic nursing training with expert knowledge and competencies, referred to by various titles (e.g. specialist nurse, public health practitioner, nurse practitioner, clinical nurse specialist, clinical nurse consultant, nurse midwife, nurse anaesthetist). Educational preparation is preferably at the master level.”*

### Phase 2: Areas in need of NARs and key barriers and challenges to developing the functions of NARs

**Delphi Round 1:** Areas in need of NARs and key barriers and challenges to developing NAR roles in each country

A total of 17 experts from 10 countries participated in round 1. The first part of round 1, i.e., areas in need of NAR within their own country identified emergency care (4.82 ± 0.53), mental healthcare (4.76 ± 0.44), clinical leadership (4.65 ± 0.86), and rural and/or remote areas (4.41 ± 1.06) as top essential areas for NAR, as noted by the highest scoring responses in each domain.

To identify differences in areas in need of NARs according to national income level, we grouped the participating counties according to the World Bank database (World Bank database, 2018), with low-income and lower-middle-income economies grouped together as low-income countries, and upper-middle-income and high-income economies categorized as high-income countries. In the Delphi round 1 of this study, high-income countries included Australia, China, Hong Kong, Japan, and South Korea; low-income countries included the Philippines, Vanuatu, and the Solomon Islands. Areas in need of NARs showed differences between the two economic levels. There was a relatively high demand for chronic disease care, nursing home care, elderly health, and mental healthcare in high-income countries, whereas all other items (e.g., emergency care, critical care, and maternal health/midwifery) were in high need in the low-income countries.

In the second part of round 1, the positive and negative factors contributing to the establishment/development of NAR roles were examined using the SWOT framework. The most highly scored strengths were accessible education and training systems (4.06 ± 1.03) and effort of national/state nurses association (4.06 ± 1.14). The most highly scored weaknesses were absence or ambiguity of a systematic career path for NARs (4.00 ± 1.12) and lack of funding sources within nursing to pursue advanced role education (3.94 ± 1.09). While respondents viewed changing demand for care (4.59 ± 0.71) and general definition of the scope of practice of NARs in legislation (4.41 ± 0.87) as possible opportunities, they noted the lack of regulations specifying roles (4.41 ± 0.94), and lack of positions available/created for NARs (4.41 ± 0.71) as threats.

The highest scoring items for how NAR roles can contribute to UHC were in the following order: Providing effective, responsive individual and population-based services, ensuring accessibility and availability (4.88 ± 0.33), participating in partnerships for public policy (4.76 ± 0.56), advising policy on incentives for appropriate provision and use of services (4.65 ± 0.61), ensuring service coverage and access (4.59 ± 0.62), advising policy on public health preparedness (4.59 ± 0.62), and advocating for health system adaptability and sustainability (4.59 ± 0.62).

**Delphi Rounds 2 & 3:** Areas in need of NARs and key barriers and challenges to developing the NAR roles in the WPR as a whole

Experts from 14 countries participated in round 2 (n=23) and round 3 (n=19) Delphi surveys. Within part 1 (areas in need of NARs) emergency care was identified as the highest need within the field-specific domain for the WPR. In the client group-specific domain, round 2 showed higher demand for maternal health/midwifery, while round 3 had lower scores and rankings. In Round 3 the highest scores were for elderly health. Perception of NAR within the WPR also found “rural and/or remote areas” as the highest area of need in the area-specific domain and “clinical leadership” as the highest area in the professional role domain (Table 2).

**Table 2. Key findings and messages of the Delphi rounds 2 & 3**

For part 2 (SWOT questions), regarding the positive and negative influences in developing NAR roles, “accessible education and training system (4.74 ± 0.56)” was the highest scoring strength, and “weak or absent leadership and advocacy by professional nursing organisation (4.42 ± 0.84)” was the highest scoring weakness. “Changing demand for care (4.58 ± 0.83)” was the highest scoring opportunity factor, and “lack of regulation specifying roles (4.58 ± 0.77)” was the highest scoring threat. Considering both the positive and negative aspects of the internal and external environments identified through SWOT analysis, strategies for developing NAR roles were prioritized (Table 2).

For part 3, which assessed the perceived need for NAR roles to assist in the goal of UHC, a high score distribution of 4 or greater overall was noted, with “advising policy on incentives for appropriate provision and use of services (4.11 ± 0.94)” and “advising policy on financial protection (4.16 ± 0.83)” having the lowest relative scores (Table 2).

### Phase 3: Policy recommendations to increase the use of NARs to improve access to health services in the WPR – Interview findings

Content analysis was performed according to the existence of NP-related regulation by country as well as by inductively identifying domain areas. The two focus areas of content analysis were: 1) areas in which NARs were currently contributing to improve access to healthcare in each country and how they can contribute in the future, and 2) strategies for increasing the use of NARs in each country as well as within the WPR at large.

We identified that NARs provide various healthcare services, such as operating independent clinics, providing patient-centred care, and responding promptly to patients, resulting in overall healthcare improvement. At the country level, eight main content areas were identified as strategies for the development of NAR roles in each country: Education/training, research, multidisciplinary approaches, remuneration, career advancement, creation of working positions, politics/registration, and needs assessment.

For specific strategies, depending on whether role related-regulations existed, we found that in countries with NP/APN role-related regulation in place, opinions favored “NAR roles must be recognized by other professionals and/or organisations” and “NARs need to develop specific abilities, such as policy-making, communication, and negotiation skills”. Conversely, in countries that did not have role-related regulations, statements related to increasing positions were emphasized, such as the need to expand the nursing role from the hospital to the community, creation of positions in the government and private sector, and the importance of a needs assessment to identify the clinical areas and communities where NARs are most needed.

At the WPR level, a three-level strategic framework to enhance the development of NAR roles was also identified - Micro (individual), organisational (networking), and macro (structural/governmental) levels. The micro level refers to practicable strategies within an individual nurse or nursing group, including increased opportunities for education, training, leadership/management capacity building, and conducting research. Secondly, improvement at the organizational level involved strategies within a broader group, which included clear paths of a career ladder system and building interdisciplinary collaboration. Thirdly, the macro level requires national efforts beyond the individual and organisation levels, such as increased remuneration for higher-level roles, as well as legislation and policy support for NAR roles. Furthermore, vision and support from organisations and governments, as well as conducting assessments to determine where NARs are most needed were also important strategies identified at the macro level.

Beyond the abovementioned strategies being enacted in each country, the participants emphasized efforts to develop stronger networking systems at the regional level. Recognizing the diversity within the WPR itself, specific examples included building a regional association for NARs, and strengthening collaboration between countries for more opportunities to share experiences and learn from each other.

### Phase 4: Integration of phases 2 & 3

Findings of the integration of the Delphi (Phase 2) and exploratory interviews (Phase 3) are summarized in Figure 1. Strategies derived from the Delphi phase aligned with the content analysis results of the exploratory interviews. The Delphi internal strategies were found to be parallel to the micro level and organizational level of the exploratory interview phase. The external strategies of the Delphi phase were also similar in context to the macro level of the exploratory interview phase. Taken together, five core strategic recommendation areas were extracted within the nursing domain and governmental-level recommendations for NARs promotion in WPR.

**Figure 1. Strategies to enhance the development of NARs identified from phases 2 & 3**

### Discussion and Recommendations

This study involved a three-step process, involving national experts from various fields, including healthcare professionals, policy makers, nurse leaders, and educators, across 18 countries in the WPR. The current study is significant, as it is the first multi-national study focusing on how NARs can contribute to equitable healthcare access in the WPR. The descriptive survey allowed for identifying the status of NARs - their functions, responsibilities, education levels, and qualifications - in several countries of the WPR. The Delphi survey identified key barriers and challenges faced by each country for enhancing the development of these nurses (round 1) as well as for the WPR (rounds 2 and 3). Lastly, through exploratory interviews, strategies for establishing NAR roles to improve equitable access to healthcare were identified. In this section, we discuss the significance and limitations of the study along with specific recommendations.

This study shows that NARs vary in nomenclature, functions, legislation, education level, and qualifications by each country. Similar reports have been made in prior studies on advancing nursing practice (Duffield, Gardner, Chang, & Catling-Paull, 2009; Sheer & Wong, 2008), although they do not pertain to the WPR. In particular, there were a number of countries that had no specific regulations at the national level but followed the standards set by a national nursing association or job descriptions by educational institutions or employing bodies, which is similar to the findings of a previous study that targeted advanced nurse practitioners (King, Tod, & Sanders, 2017). Although the ICN definition of NP/APN (ICN, 2008) appears to view these terms as interchangeable, to our knowledge there is current dialogue within the Council to further differentiate NP from APN, which has implications for countries that are seeking to develop a variety of advanced roles for nurses who have appropriate expertise. However, as the WPR comprises 37 member states, including many island countries, the term NARs was used very broadly in this study in order to encompass as many ‘advanced’ nursing roles as possible, i.e., roles extending beyond the basic level of nursing, so that nurses’ roles in a wider range of tasks and contexts could be explored. Even with the agreed upon working definition of NARs used in the study, however, the potential variation in interpreting the advanced role types that emerged from Phase 1 data may require caution. For example experts who participated in Phases 2 and 3 sometimes appeared to answer referring to different types of NARs even if they were from the same country of Phase 1 respondents. It is possible that the scope of responses may have varied because the perception of the meaning of NARs differed, or the experts did not recognize working areas outside of their own field and/or setting.

The areas in need of NARs were found to be different between the national and regional levels. There was a tendency for needs to be assessed as lower priority in Delphi rounds 2 & 3 when participants were asked to respond for the WPR, compared to round 1, which focused on the context of the respondent’s own country. It might be that experts responded conservatively for the region because they did not have full comprehension of the context of the diverse countries that comprise the WPR. However, as noted in Table 2, very little changes in priorities were observed in most categories between these questioning rounds.

In this study, strategies for enhancing the development of NARs were grouped into internal strategies at the micro (individual) and organisational (networking) levels, as well as external strategies that pertained to the macro (structural) level (Figure 1). In several previous studies, legislation, educational development, and credentialing have been emphasized as crucial for the development of NP/APN (King et al., 2017; Wainwright, Klein, & Daly, 2016; Wysocki, Underwood, & Kelly-Weeder, 2015). Our findings parallel the strategies reported by a prior study conducted at the national level in Canada, which explored various strategies for developing advanced nursing roles, in this case specialized NPs (McNamara, Giguere, St-Louis & Boileau, 2009). Experts in our study also emphasized additional specific strategies, such as building collaboration, developing a career structure, and exploring the professional development needs of nurses in these roles. In particular, collaboration among organisations and countries are emphasized among our study participants. Given that collaboration among nurses performing advanced roles and responsibilities improves role integration, autonomy, role clarity, and team capacity (Burgess & Purkis, 2010; Nardi & Diallo, 2014), this implication should be emphasized. Considering differences in context, such as health systems, health status, income level, and status of NARs, country-specific strategies are required as a subset of the more generic “region-wide” strategies described in this study.

UHC, which is an important sustainable development goal, means that all individuals should be able to access high-quality healthcare without financial difficulty. Study participants emphasized that NARs are well positioned to meaningfully contribute to the achievement of UHC, as evidenced in Phase 2 (Delphi), where all five UHC subcategories (quality, efficiency, equity, accountability, and sustainability/resilience) showed a relatively high mean score of four or higher. In contrast, the item “advising policy on financial protection and incentives for appropriate provision and use of services” was scored as a somewhat lower priority. This appears to suggest that the first foundational step is to establish remuneration and financially-related support systems for NAR before becoming involved in policy. It is also possible that financial leadership is easily overlooked in NARs as clinical-related issues are prioritized. Therefore, in addition to clinical expertise, developing policy savvy knowledge, political competence, and understanding of finances through training and NAR educational curricular are warranted.

This study has some limitations, one being that a greater number of experts (2-3 experts per country) of the eight countries with WHO CCs mainly contributed to the study, compared to one to two experts from ten countries. The status of other countries in the WHO WPR that were not included in this study was not addressed; thus, the findings are limited to reflect the context of all countries within the WPR. In order to overcome these limitations, however, the literature was continuously reviewed for WPR contexts and situations, and experts from six additional countries were invited (a total of 14 countries) were invited to the Delphi, to capture the overall situation within the region. Although the researchers shared English summaries after exploratory interviews were conducted in each country’s specific language, there was a limit to the richness of narratives that could be attained, possibly affecting analyses of all pertinent information.

In essence, we were able to draw five core strategic recommendation areas within the nursing domain to assist in the development of NAR roles both at the organisational level (i.e., department of nursing within a hospital) and relating to professional nursing associations. The 15 strategies presented in Table 3 specifically address improving education/training, conducting research on NARs, creating career development pathways, enlisting multidisciplinary support, and building cross-country collaboration.

**Table 3. Strategic recommendations for the development of NARs within the nursing domain**

While strategic action is needed within nursing, government-level involvement and degree of commitment will ultimately promote or hinder NAR potential contributions to the health. We present four core-level recommendations for policy at the governmental-level, highlighted in Figure 2. If governments recognize the potential of this sizable health professional group, action is required to take a foundational step is establishing legislation on NAR and their scope of practice, and creating a professional regulatory system. Second, appropriate remuneration measures for NAR that align with career structures will strengthen the infrastructure for NAR development and sustainability. Third, establishing supportive channels within government and private sectors and improving public understanding of NAR through media and social network platforms has the power to increase NAR involvement in the community and at the policy table. Finally, as rapid shifts in health issues and demographics are increasingly being observed, comprehensive national needs assessments are required to carefully examine priority areas for NAR, which can then guide educational preparation at the national-level (Benner, 2012).

**Figure 2. Governmental-level recommendations for the promotion of NARs**

## Conclusions

We found that demand for NARs is high in the WPR and presented recommendations for both the nursing community and at governmental-level to enhance NAR development and contributions for equitable healthcare access. We suggest further research that collates and highlights best practices for how NAR development and collaboration are best realized within diverse sociocultural contexts, especially in relation to UHC at local and regional levels. Also research that coordinates dialogue among nurses in clinical and/or policy positions, academia, and international organizations such as ICN, is needed to clarify advanced nursing roles across NP, APN, and NAR.

### Clinical Resources

International Council of Nurses. (2016). Advanced Practice Nursing: An essential component of country level human resources for health. https://www.who.int/workforcealliance/knowledge/resources/ICN_PolicyBrief6AdvancedPracticeNursing.pdf?ua=1 United Nations. (2015). Transforming our world: The 2030 agenda for Sustainable Development. https://www.un.org/ga/search/view_doc.asp?symbol=A/RES/70/1&Lang=E/

## Data Availability

The data that support the findings of this study are available from the corresponding author upon reasonable request.

## Acknowledgements

This study was funded by the Yonsei University College of Nursing (grant number: 6-2017-0112).

We would like to thank the following colleagues for their assistance with the sampling and collection of data: Caryn West, Erika Ota, Gyungjoo Lee, Peter James Abad, Michael Larui, Amelia Fuha’amago, and Silina R Waqa.

## Declarations of interest

none

---

**Table S1.**
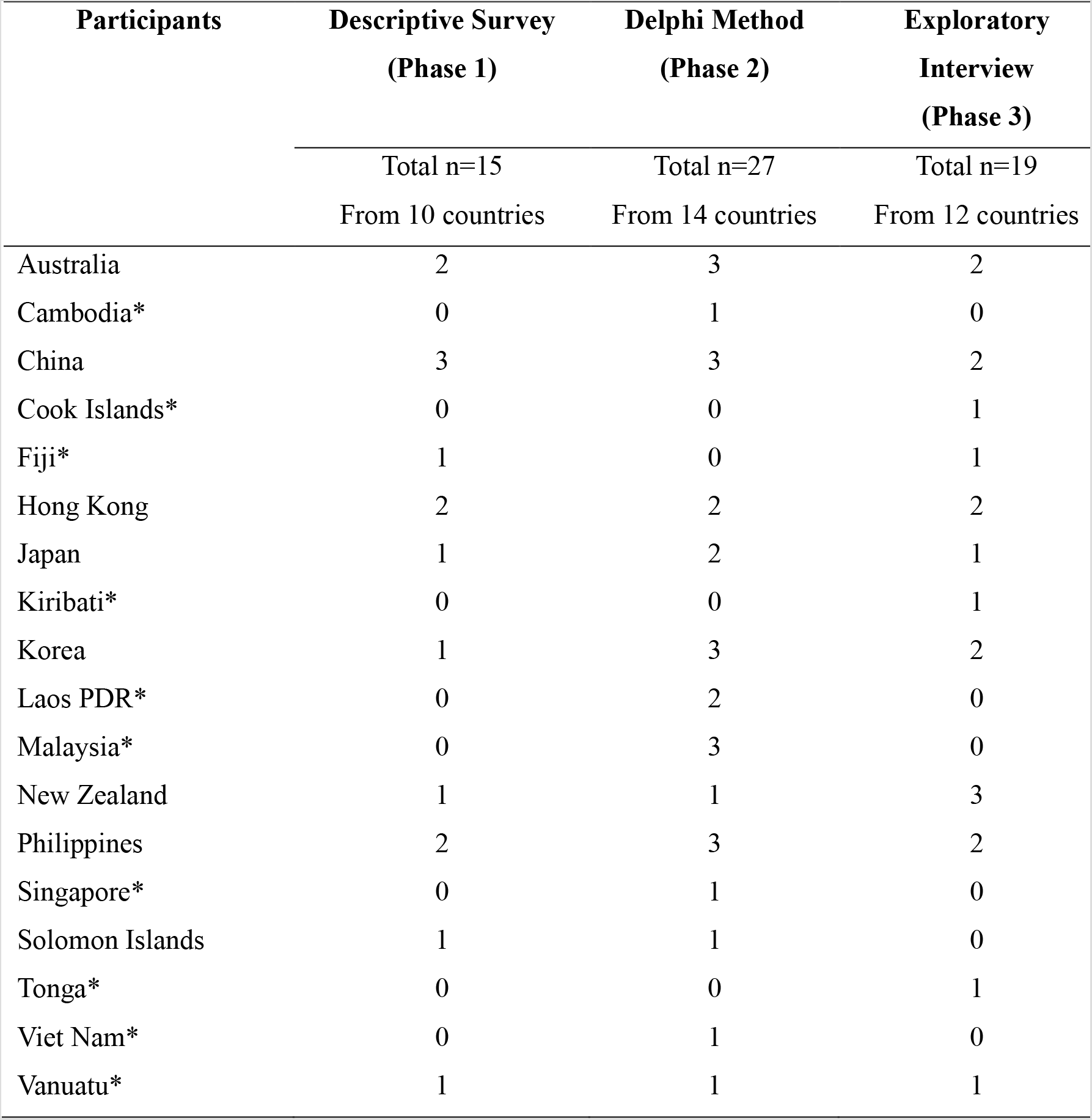
Participants in each phase

**Table S2.** Key findings and messages of the Delphi rounds 2 & 3

| **Part 1: Fields Where NAR Are Needed** |  |
| --- | --- |
| Domain | Specific Area |
| Field-Specific | Emergency care |
|  | Critical care |
| Client group-specific | Elderly health |
|  | Child health |
|  | Rural and/or remote area |
|  | Clinical leadership |
|  | Professional leadership |
| **Part 2: Strategies for NAR Development** |  |
| Level | Strategy |
| Internal | Highlight the need of NAR and demand for care through policy (SO strategy) |
|  | Need input to develop leadership and advocacy by professional nursing organization (WO strategy) |
|  | Establish career ladder system and specific position and education/training system (WO strategy) |
|  | Identify current contribution of NARs and the future need (ST strategy) |
| External | Access the required NAR role in the context, then match the NAR who are already qualified (SO strategy) |
|  | Establish regulation specifying roles and scope of practice (ST strategy) |
|  | Legislate the education and training system as well as qualification (WT strategy) |
| **Part 3: NAR Role to Achieve UHC** |  |
| UHC Category | NAR Role |
| Quality | Providing effective, responsive individual and population-based services ensuring accessibility and availability |
|  | Practicing individual, family, and community engagement |
| Accountability | Voicing to influence government leadership and rule of law for health |
|  | Participating in partnership for public policy |
| Sustainability & Resilience | Advocating for health system adaptability and sustainability |
|  | Promoting/engaging in community capacity development |

NAR = Nurses in Advanced Roles; UHC = Universal Health Coverage; SO = Strength-Opportunity; WO = Weakness-Opportunity; ST = Strength-Threat; WT = Weakness-Threat

**Table S3.** Strategic recommendations for the development of NARs within the nursing domain

| **Domain** | **Strategic recommendations** |
| --- | --- |
| **Education & Training** | 1. Master's degree (or a doctorate) is recommended<br>2. Create a structured program through a nursing school or training institutions<br>3. Train for specific abilities (e.g., decision-making, leadership, budgeting, negotiation skills, etc.)<br>* This is also expected to contribute to achieve UHC<br>4. Offer training in other countries or visiting trainers from developed countries if resources are insufficient |
| **Research** | 1. Facilitate research about evidence-based, clinically-based issues<br>2. Conduct research about NARs to evaluate efficacy and highlight improvements in clinical outcomes. |
| **Career Development** | 1. Clarify career progression (e.g., career ladder system) of nurses at organizational level<br>2. Create pathways for nurses to go into various positions (e.g., health administration, primary healthcare, government, private sector, academia, etc.)<sup>a</sup> |
| **Multidisciplinary Approach** | 1. Strengthen multidisciplinary teams (including nurses, physicians, and other health providers) or cross-cluster meetings from planning to decision-making<br>2. Build recognition of NAR from other professionals and organizations; Identify supportive sources from organizations and government<sup>a</sup> |
| **Collaboration across Countries** | 1. Identify and link with support for NARs at regional level<br>2. Develop collaborative efforts between countries to share experiences and learn from each other (e.g., developing programs and practice)<br>3. Create an association for NARs regionally as well as globally |

<sup>a</sup> If the country does not have NAR role related regulation.

**Figure S1.**
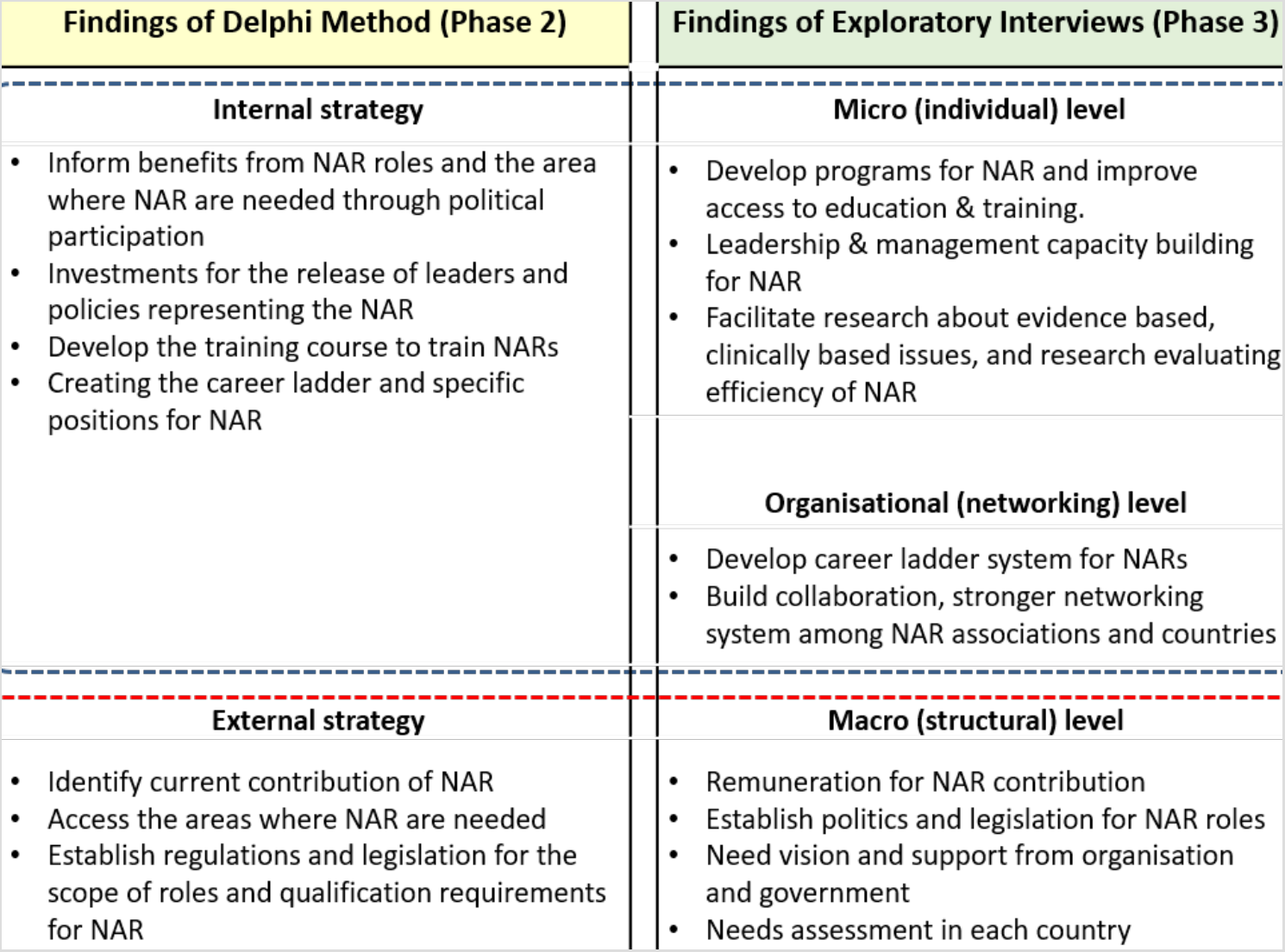
Strategies to enhance the development of NARs identified from phases 2 & 3

**Figure S2.**
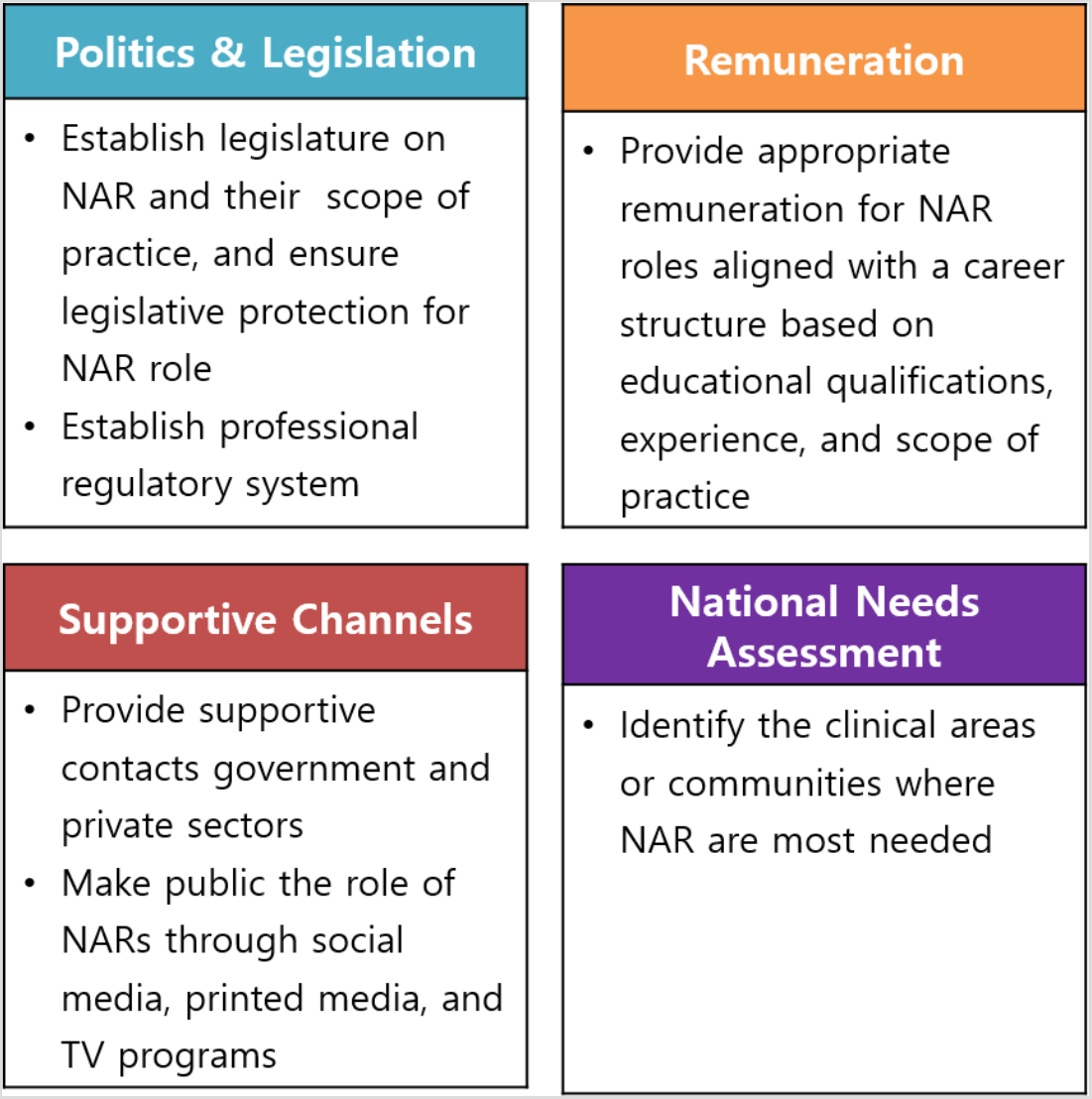
Governmental-level recommendations for the promotion of NARs

## Notes

### Competing Interest Statement

The authors have declared no competing interest.

